# Smoking and subfertility: multivariable regression and Mendelian randomization analyses in the Norwegian Mother, Father and Child Cohort Study

**DOI:** 10.1101/2021.10.25.21265469

**Authors:** Álvaro Hernáez, Robyn E. Wootton, Christian M. Page, Karoline H. Skåra, Abigail Fraser, Tormod Rogne, Per Magnus, Pål R. Njølstad, Ole A. Andreassen, Stephen Burgess, Deborah A. Lawlor, Maria Christine Magnus

## Abstract

**Objective:** To investigate the association between smoking-related traits and subfertility.

**Design:** Prospective study.

**Setting:** Nationwide cohort in Norway.

**Patients:** 28,606 women (average age 30) and 27,096 men (average age 33) with questionnaire and genotype information from the Norwegian Mother, Father and Child Cohort Study.

**Intervention:** Self-reported information on smoking (having ever smoked [both sexes], age at smoking initiation [women only], smoking cessation [women only], and cigarettes smoked per week in current smokers [both sexes]) was gathered. Genetically predetermined levels or likelihood of presenting the mentioned traits were estimated for Mendelian randomization (MR) analyses.

**Main outcome measure:** Subfertility, defined as time-to-pregnancy ≥12 months.

**Results:** A total of 10% of couples were subfertile. In multivariable regression accounting for age, years of education, body mass index, and number of previous pregnancies, having ever smoked was not linked to subfertility in women or men. A higher intensity of tobacco use in women who were current smokers was related to greater odds of subfertility (+ 1 standard deviation [SD, 48 cigarettes/week]: odds ratio [OR] 1.12, 95% confidence interval [CI] 1.03 to 1.21), also after adjusting for the partner’s tobacco use. Later smoking initiation (+ 1 SD [3.2 years]: OR 0.89, 95% CI 0.84 to 0.95) and smoking cessation (relative to not quitting: OR 0.83, 95% CI 0.75 to 0.93) were linked to decreased subfertility in women who had ever smoked. Nevertheless, MR results were not directionally consistent for smoking intensity and cessation and were imprecisely estimated in two-sample MR, with wide confidence intervals that overlapped with the multivariable regression results. In men, greater smoking intensity was marginally linked to greater odds of subfertility in multivariable analyses, but this association was attenuated when adjusting for the partner’s smoking intensity (+ 1 SD [54 cigarettes/week]: OR 1.05, 95% CI 0.96 to 1.15). MR estimates were directionally consistent but again imprecisely estimated.

**Conclusions:** We did not find robust evidence of an effect of smoking on subfertility. This may be due to a true lack of effect, weak genetic instruments, or other kinds of confounding. The relevant limitations across all methods highlights the need for larger studies with information on subfertility.

## INTRODUCTION

Smoking is a well-known source of thousands of pro-oxidative and pro-inflammatory compounds (1), capable of damaging reproductive tissues which in turn may compromise fecundity (2-4). Observational studies in women have reported that active smoking was linked to 14% greater odds of subfertility (trying to conceive for ≥12 months) (5) and smoking intensity was dose-dependently associated with greater subfertility risk (6). Tobacco use has also been related to surrogate indicators of decreased fertility such as accelerated follicular depletion and earlier menopause (7, 8). Although smoking has been linked to oligozoospermia and morphological defects of sperm (9), two prospective studies reported no association between tobacco use and subfertility in men (10, 11). Considering this evidence, the Practice Committee of the American Society of Reproductive Medicine has suggested a causal effect of tobacco use on clinical subfertility (12). However, existing evidence is subject to numerous methodological limitations. The primary concerns raised were that the relationship between smoking and decreased fertility has not been shown to be sufficiently strong, residual confounding cannot be ruled out, and most studies were retrospective and thus unable to reveal any potential exposure-to-effect sequence (12).

The use of complementary methodological approaches with different strengths and sources of bias could help clarify whether there is a causal relationship between tobacco use and subfertility (13). Mendelian randomization (MR) is based on the use of genetic variants that are linked to an exposure (e.g. having ever smoked, a greater intensity of current tobacco use) to assess the unconfounded effect of this exposure on a certain outcome (e.g. subfertility) (14). It can be performed using data from a single sample (one-sample MR: exposure, outcome, and genetic variants are measured in the same population) or from two samples (two-sample MR: the association between genetic variants and the exposure are assessed in one population and the association between genetic variants and the outcome are studied in a second population) (15). In this paper, we compare results from multivariable logistic regression, one-sample MR, and two-sample MR, considering that these are affected by different and unrelated sources of bias when studying the association between smoking-related traits and subfertility (15-17) (**Table 1**). Similar results in all of them would allow for more robust conclusions (13).

**Table 1.** Comparison among multivariable logistic regression, one- and two-sample Mendelian randomization regarding sources of bias in the association between smoking and subfertility

|  | **Residual confounding**<br>(the effect of smoking on subfertility could be due to behavioral or socioeconomic conditions intimately related to smoking, such as poor diet, poorer health status, etc.) | **Weak genetic instruments**<br>(the available genetic instruments for smoking traits explain a small proportion of the variability of the exposure) | **Horizontal pleiotropy**<br>(the genetic instruments for smoking traits influence the risk of subfertility via mechanisms other than smoking) | **Confounding of the genetic instrument-outcome association**<br>(by population stratification) |
| --- | --- | --- | --- | --- |
| Multivariable regression | High risk | Unaffected | Unaffected | Unaffected |
| One-sample MR | Low risk | Overestimated associations.<br>Consistence with two-sample MR estimates minimizes the risk of this bias | Possibility to explore the association between genetic instruments and other subfertility risk factors.<br>Multivariable MR could then be used to correct for such bias | Adjustment for ancestry-informative principal components minimizes the risk of this bias |
| Two-sample<br>MR | Low risk | Bias towards the null association.<br>The Robust Adjusted Profile Score<br>approach is immune to this bias | Consistence among MR methods,<br>lack of between SNP heterogeneity,<br>and lack of outliers in MR scatterplots<br>minimizes the risk of this bias | High risk |

Our aim was to investigate the association between tobacco use and subfertility in women and men by multivariable logistic regression and one- and two-sample MR.

## METHODS

### Study participants

Our study used data from the Mother, Father and Child Cohort Study (MoBa) (18, 19). The MoBa Study is a population-based pregnancy cohort study conducted by the Norwegian Institute of Public Health. Pregnant women and their partners across Norway were recruited between 1999 and 2008 at the time of the routine ultrasound screening (around 18^th^ gestational week). The cohort now includes 114,500 children, 95,200 mothers and 75,200 fathers. The current study is based on version #12 of the quality-assured data.

For the current study, we defined a subsample of parents with available genotype data and pre-pregnancy information on tobacco use. The genotype data in this study came from blood samples gathered from both parents during pregnancy (20) and followed the pipeline described by Helgeland et al (genotype calling, imputation, and quality control) (21). Our work is described according to the STROBE guidelines for reporting MR and cohort studies (22, 23).

### Tobacco use

Parents responded to questions related to their smoking habits in the questionnaire completed at recruitment. First, they reported if they had ever smoked. After an affirmative answer, participants reported their age when they started smoking, if they were smokers when they conceived, if they had quit smoking (and the date when they quit), and the amount of cigarettes per week they currently smoked (or used to smoke for former smokers). Using these data, we defined three exposure variables: ever smoker (yes/no, available for the whole population), age at smoking initiation (continuous, available in current/former smokers), and smoking cessation (yes/no, available in current/former smokers). In addition, we computed the average cigarettes smoked per week at conception or during the two years prior to conception (continuous, available in current smokers and in participants who smoked at this time).

### Selection of genetic variants and generation of instrumental variables

Genetic instruments were extracted from the most recent genome-wide association study (GWAS) on smoking-related traits (24). It included more than 1.2 million individuals (none of them participated in the MoBa cohort) and reported 378 conditionally independent single nucleotide polymorphisms (SNPs) associated with smoking initiation, 10 linked to the age of smoking initiation, 24 related to smoking cessation, and 55 associated with smoking heaviness (cigarettes per week) (24). Independent SNPs were defined according to linkage disequilibrium blocks across the genome using the methods by Li J et al., Gao X et al., and Chen Z et al. (25-27), presented a minor allele frequency ≥ 0.1% and were associated with their respective phenotypes according to the standard genome-wide significance threshold (*p* < 5 × 10^−8^). A total of 355, 10, 23, and 50 independent SNPs were available for smoking initiation, age of smoking initiation, smoking cessation, and smoking heaviness, respectively, in the MoBa genotype database.

For the one-sample MR analyses, we generated a weighted genetic risk score (GRS) by multiplying the number of risk alleles by the effect estimate of each variant and dividing this value by the total number of SNPs (28). SNPs were used individually as the genetic instruments in the two-sample MR.

### Subfertility

Women were asked at recruitment if the pregnancy was planned and to provide information on how many months it had taken them to conceive (19). The answer options were “<1 month”, “1-2 months” and “3 or more months”. If the mother responded “3 or more months”, she was asked to further specify how many months they were trying to conceive. We defined subfertility as trying to conceive for ≥12 months or having used assisted reproductive technologies. Those reporting trying for <12 months (defined as fertile) were included in the reference group. Participants involved in unplanned pregnancies were excluded.

### Other variables

Information on the age of the participants (continuous), educational level (years of education equivalent to the US system (29, 30), continuous), pre-pregnancy body mass index (BMI, continuous), and previous number of deliveries (0, 1, 2, or ≥3) was gathered in the baseline visit of the study as potential confounders.

### Ethical approval

The MoBa cohort is conducted according to the Declaration of Helsinki for Medical Research involving Human Subjects. The establishment of MoBa and initial data collection was based on a license from the Norwegian Data Protection Agency. It is now based on regulations related to the Norwegian Health Registry Act. Participants provided a written informed consent before joining the cohort. This project was approved by the Regional Committee for Medical and Health Research Ethics of South/East Norway (reference: 2017/1362).

### Statistical analyses

Normally distributed continuous variables were described by means and standard deviations (SD), non-normally distributed continuous variables by medians and 25^th^-75^th^ percentiles, and categorical variables by proportions. Differences in baseline characteristics between subfertile and fertile parents, and between participants with and without genotype information in the MoBa cohort, were investigated by t-tests in normally distributed continuous variables, Mann-Whitney U-tests in non-normally distributed continuous variables, and chi-squared tests in categorical variables.

#### Multivariable regression analyses

We assessed the association between smoking-related traits and subfertility in women and men by multivariable logistic regressions (adjusted for predefined subfertility risk factors: age, years of education, BMI, and number of previous pregnancies (31, 32)). For binary exposures (smoking initiation and smoking cessation), we investigated the differences in the odds of subfertility in those exposed compared with those who were not. For continuous variables, we assessed the relationship between an increase in 1 SD in age at smoking initiation (in ever smokers) and the number of cigarettes smoked per week (in current smokers) with odds of subfertility. We also assessed whether a model using smoothed cubic splines (*K*+4 degrees of freedom) fitted the data better than a linear function using a likelihood ratio test. In those traits in which there was information from both parents (smoking initiation and number of cigarettes smoked per week), we further adjusted logistic regression models for the partner’s trait to minimize bias due to assortative mating. Clustered standard errors were computed to account for non-independence between pregnancies in participants with data on multiple pregnancies.

#### One-sample Mendelian randomization

We used logistic regression to estimate the genetically predicted likelihood of smoking initiation (in both sexes) and smoking cessation (in women), and linear regression to calculate the genetically predicted values of age at smoking initiation (in women) and the number of cigarettes smoked per week (in both sexes), using their respective GRSs as predictors. Next, we assessed the linear relationship between an increase in one SD in the genetically predicted traits and subfertility using logistic regression.

#### Two-sample Mendelian randomization

We first performed two GWASs (one for women and one for men) to obtain summary associations of each SNP, across the genome, with subfertility in the MoBa cohort. Full details of this procedure are available in the **Supplemental Materials**. Next, in these subfertility GWAS summary data, we looked for the SNPs related to each of the smoking traits and extracted the information related to their association with subfertility. We harmonized both datasets and excluded palindromic SNPs with minor allele frequencies close to 0.5 (24, 33). Following these exclusions, we had 301, 43, 7 and 16 SNPs for analyses of smoking initiation, intensity, age at initiation, and cessation, respectively. Finally, we used inverse variance weighted regression as the main two-sample MR analysis (15).

#### Verification of MR assumptions

The key assumptions of MR are: 1) the genetic instrument is robustly related to the exposure, 2) the genetic instrument is only associated with the outcome through the exposure of interest, and 3) there is no confounding of the genetic instrument-outcome associations (15).

Regarding the first assumption, we checked the strength of the association between the genetic instruments and their phenotypes. For binary exposures we used logistic regression, area under the ROC curve, and pseudo-*R*^*2*^ by the Nagelkerke method, and for continuous exposures we used linear regressions, *F*-statistics and *R*^*2*^. Since weak instruments deviate MR causal estimates towards the null in two-sample MR, concordance between one-sample and two-sample MR reduces the concern that estimates might be influenced by weak instrument bias (34). We also performed the Robust Adjusted Profile Score two-sample MR method, which is unbiased by weak instruments (34, 35). The second assumption may be violated when the genetic instruments influence other risk factors for the outcome independently of the exposure of interest (horizontal pleiotropy) (16). To evaluate horizontal pleiotropy in one-sample MR, we checked the association of GRSs with known risk factors for subfertility. We studied the relationship between 1 SD increase in the GRS and the risk factors (age, years of education, BMI, number of previous deliveries) (31, 32) using linear regressions. If any of the GRSs was associated with a risk factor, we considered that a potential pleiotropic effect was present for all the smoking-related traits. We then performed multivariable MR analyses if GWAS data for the potential pleiotropic variable was available (17). There was evidence of one or more of the smoking trait GRS associating with education and BMI and we were able to undertake multivariable MR for both. For education we used the GWAS by Lee JJ et al. (*n* = 1,271 independent SNPs, ∼1.1 million participants (36); 1,159 of the SNPs were available in MoBa). For BMI we used the GWAS by Yengo L et al (*n* = 941 independent SNPs, ∼700.000 participants (37); 896 of the SNPs were available in MoBa). We generated GRS for education and BMI using the same method as used for the smoking traits and then included the GRS for education and BMI in the one-sample MR regression models. (17). The genetic smoking instruments also associated with age, and we performed stratified analyses according to age (below vs. over the median). In addition, we estimated the association between the GRSs for smoking traits and subfertility in non-exposed participants (the GRSs for age of smoking initiation or smoking cessation in never smokers, and the GRS for current smoking intensity in never + former smokers). As we would expect no association in non-exposed participants, any evidence of one would indicate the presence of bias.

In two-sample MR we explored unbalanced horizontal pleiotropy through sensitivity analyses comparing the main estimates to those obtained from MR-Egger, the weighted median and weighted mode methods (38-40). The inverse variance weighted method assumes no unbalanced horizontal pleiotropy as it forces the regression line through SNP-exposure and SNP-outcome coordinates to go through zero. MR-Egger does not make this assumption and it does not force the line through zero. A non-zero intercept is an indication of unbalanced horizontal pleiotropy and the slope is subsequently corrected. The weighted median and weighted mode analyses are valid if less than 50% of the weight comes from SNPs that are not related to other risk factors for the outcome. Concordance in the estimates across the different approaches reduces the concern regarding unbalanced pleiotropy (38-40). We also checked for influential outliers in the variant-specific causal estimates (indicative of horizontal pleiotropy) in scatterplots (38-40). Finally, we evaluated between SNP heterogeneity, using Cochran’s *Q* and the Rücker’s *Q’* with the inverse variance weighted and Egger regression methods, respectively. Heterogeneity indicates a possible violation of the MR assumptions, of which pleiotropy is probably a major cause (38-40).

To reduce the potential for confounding of the genetic instrument-outcome association due to population stratification (third assumption), we adjusted for the first 10 ancestry-informative principal components in the one-sample MR (41).

#### Software

Analyses were performed in R Software version 4.0.3 (packages: *compareGroups, estimatr, ggplot2, miceadds*, and *TwoSampleMR*) and the GWASs to determine which SNPs were associated with subfertility in the MoBa cohort in Plink v1.9 and GWAMA (42, 43). Code for data management and statistical analysis has been made available in https://github.com/alvarohernaez/MR_smoking_subfertility_MoBa/.

## RESULTS

### Description of the study population and genetic instruments

Our study population consisted of 28,606 women and 27,096 men with genotype information (**Figure 1**). A total of 10% of the couples were subfertile. Women and men who were subfertile were older, had a lower educational attainment, had a higher BMI, and were more likely to be pregnant for the first time. In relation to smoking habits, there was a higher proportion of ever smokers in subfertile couples and, among tobacco users, smoking intensity was higher in subfertile individuals (**Table 2**). MoBa participants without genotype data were not meaningfully different to those with genetic data in age, years of education, BMI, or number of previous pregnancies compared to those with genotype data. However, participants with genotype data were more likely to be subfertile, presented a different proportion of ever smokers (women were less likely and men were more prone to have ever smoked), and showed a lower smoking intensity among current smokers (**Supplemental Table 1**).

**Table 2.** Population characteristics among genotyped participants

|  | Women |  |  |  | Men |  |  |  |
| --- | --- | --- | --- | --- | --- | --- | --- | --- |
|  | All<br>(n = 28,606) | Subfertility<br>reported<br>(n = 3,439) | No subfertility<br>reported<br>(n = 25,167) | p-value | All<br>(n = 27,096) | Subfertility<br>reported<br>(n = 3,275) | No subfertility<br>reported<br>(n = 23,821) | p-value |
| Age (years),<br>mean ± SD | 30.3 ± 4.15 | 31.5 ± 4.36 | 30.2 ± 4.09 | <0.001 | 32.7 ± 4.90 | 34.1 ± 5.36 | 32.6 ± 4.81 | <0.001 |
| Education years,<br>mean ± SD | 17.5 ± 3.11 | 17.0 ± 3.33 | 17.6 ± 3.08 | <0.001 | 16.6 ± 3.50 | 16.2 ± 3.54 | 16.6 ± 3.49 | <0.001 |
| BMI (kg/m<sup>2</sup>),<br>median (25<sup>th</sup>-75<sup>th</sup> percentiles) | 23.1<br>(21.2-25.9) | 23.7<br>(21.5-27.2) | 23.1<br>(21.1-25.7) | <0.001 | 25.5<br>(23.7-27.7) | 25.8<br>(24.0-28.1) | 25.4<br>(23.7-27.7) | <0.001 |
| Previous pregnancies<br>n (%): |  |  |  | <0.001 |  |  |  | <0.001 |
| 0 | 12,888<br>(45.1%) | 2,020<br>(58.8%) | 10,868<br>(43.2%) |  | 12,282<br>(45.4%) | 1,923<br>(58.8%) | 10,359<br>(43.5%) |  |
| 1 or more | 15,680<br>(54.9%) | 1,415<br>(41.2%) | 14,265<br>(56.8%) |  | 14,784<br>(54.6%) | 1,348<br>(41.2%) | 13,436<br>(56.5%) |  |
| Ever smokers (all<br>participants), n (%): | 13,389<br>(46.8%) | 1,722<br>(50.1%) | 11,667<br>(46.4%) | <0.001 | 13,186<br>(48.7%) | 1,668<br>(50.9%) | 11,518<br>(48.4%) | 0.006 |
| Age of smoking initiation<br>(current + former smokers),<br>median (25<sup>th</sup>-75<sup>th</sup> percentiles) | 17.0<br>(15.0-19.0) | 16.0<br>(15.0-18.0) | 17.0<br>(15.0-19.0) | 0.017 | - | - | - | - |
| Current + former smokers<br>who quit smoking, n (%): | 7,627<br>(57.0%) | 910<br>(52.8%) | 6,717<br>(57.6%) | <0.001 | - | - | - | - |
| Cigarettes/week<br>(current smokers),<br>median (25<sup>th</sup>-75<sup>th</sup> percentiles) |  | 42.0<br>(10.0-70.0) | 52.5<br>(12.0-105) | 40.0<br>(10.0-70.0) | <0.001 | 70.0<br>(21.0-105) | 70.0<br>(35.0-105) | 70.0<br>(21.0-105) |

**Figure 1.**
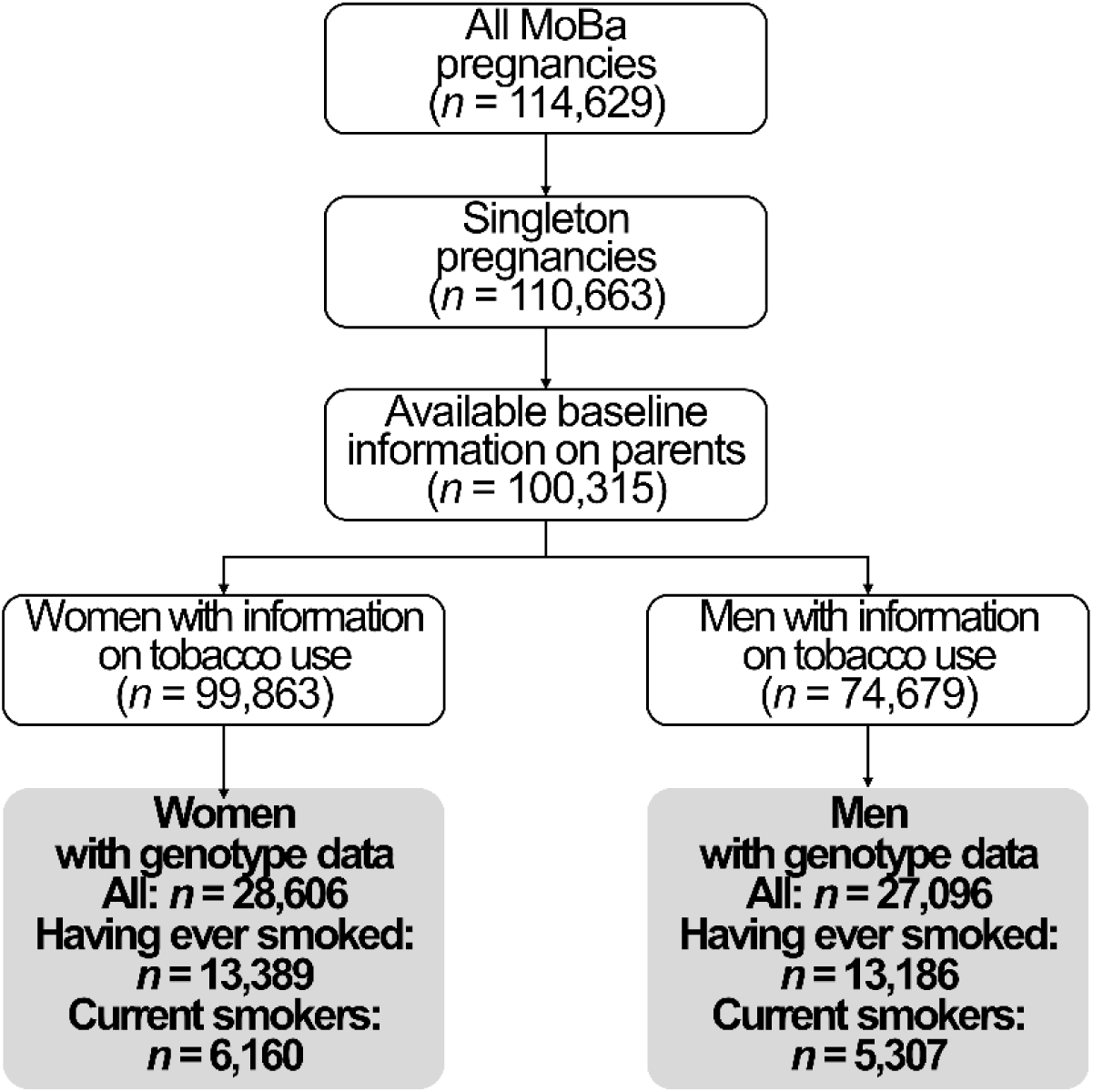
Study flow chart

**Table 3** shows the associations of the GRS for each trait with smoking traits in MoBa together with statistics related to instrument strengths.

**Table 3.** Description of genetic instrumental variables^1^

| **Binary exposures** |  |  |  |  |  |
| --- | --- | --- | --- | --- | --- |
|  | SNPs<br>(*n*) | GRS value<br>(mean ± SD) | Likelihood of presenting the exposure<br>per each 1-point increase in the GRS<br>OR (95% CI) | Pseudo-*R*<sup>2</sup> | Area under<br>the ROC<br>curve |
| GRS for smoking<br>initiation (women) | 355 | 375 ± 11.9 | 1.02 (1.018 to 1.022) | 1.87% | 0.57 |
| GRS for smoking<br>initiation (men) | 355 | 376 ± 11.8 | 1.02 (1.017 to 1.022) | 1.69% | 0.56 |
| GRS for smoking<br>cessation (women) | 23 | 24.5 ± 2.52 | 1.03 (1.019 to 1.047) | 0.22% | 0.52 |
| **Continuous exposures** |  |  |  |  |  |
|  | SNPs<br>(*n*) | GRS value<br>(mean ± SD) | Change in the exposure<br>per each 1-point increase in the GRS<br>Coef. (95% CI) | *R*<sup>2</sup> | *F*-statistic |
| GRS for cigarettes<br>smoked per week (women) | 50 | 51.2 ± 6.90 | 0.80 (0.62 to 0.98) | 1.43% | 76 |
| GRS for cigarettes<br>smoked per week (men) | 50 | 51.2 ± 6.96 | 0.87 (0.66 to 1.09) | 1.34% | 64 |
| GRS for age of smoking<br>initiation (women) | 10 | 7.78 ± 1.83 | 0.023 (-0.014 to 0.059) | 0.01% | 1 |

1. The associations of smoking cessation and age of smoking initiation with their
respective GRSs in men could not be validated due to the lack of information on this
phenotype.

### Comparison of main multivariable regression, one-sample, and two-sample MR results

**Figure 2A** shows the confounder-adjusted associations, one-sample, and two-sample MR results for each smoking trait in women. For smoking initiation, there were similar close to the null results in both multivariable regression (odds ratio [OR] 1.03, 95% confidence interval [CI] 0.95 to 1.11) and one-sample MR (OR 1.01, 95% CI 0.98 to 1.05), and an inverse, but also close to the null, relationship in two-sample MR (OR 0.90, 95% CI 0.75 to 1.09). Regarding smoking intensity, it was linearly linked to greater odds of subfertility in multivariable regression (1 SD increase in the number of cigarettes smoked per week [+48 cigarettes/week]: OR 1.12, 95% CI 1.03 to 1.21; *p*-value_non-linearity_ = 0.970), even after adjusting for the partner’s smoking heaviness (OR 1.10, 95% CI 1.01 to 1.19; **Supplemental Table 2**). However, close to the null results were observed in one-sample MR (OR 0.96, 95% CI 0.89 to 1.03) and a notable inverse, but imprecisely estimated result in two-sample MR analyses (OR 0.68, 95% CI 0.40 to 1.15). For age at smoking initiation, a broadly similar pattern of results across the three measures was seen. We observed a linear association between later smoking initiation and lower odds of subfertility in multivariable analyses (1 SD increase in the age of smoking initiation in ever smokers [+3.2 years]: OR 0.89, 95% CI 0.84 to 0.95; *p*-value_non-linearity_ = 0.933), close to the null results in one-sample MR (OR 0.96, 95% CI 0.91 to 1.02), and a greater, directionally concordant but imprecise association in two-sample MR (OR 0.47, 95% CI 0.11 to 2.01). Finally, for cessation amongst those who had ever smoked, there were similar, in terms of OR, inverse associations in multivariable regression (OR 0.83, 95% CI 0.75 to 0.93) and two-sample MR (OR 0.80, 95% CI 0.46 to 1.38), the latter with wide confidence intervals, whereas the result in one-sample MR was close to the null (OR 1.04, 95% CI 0.99 to 1.09).

**Figure 2.**
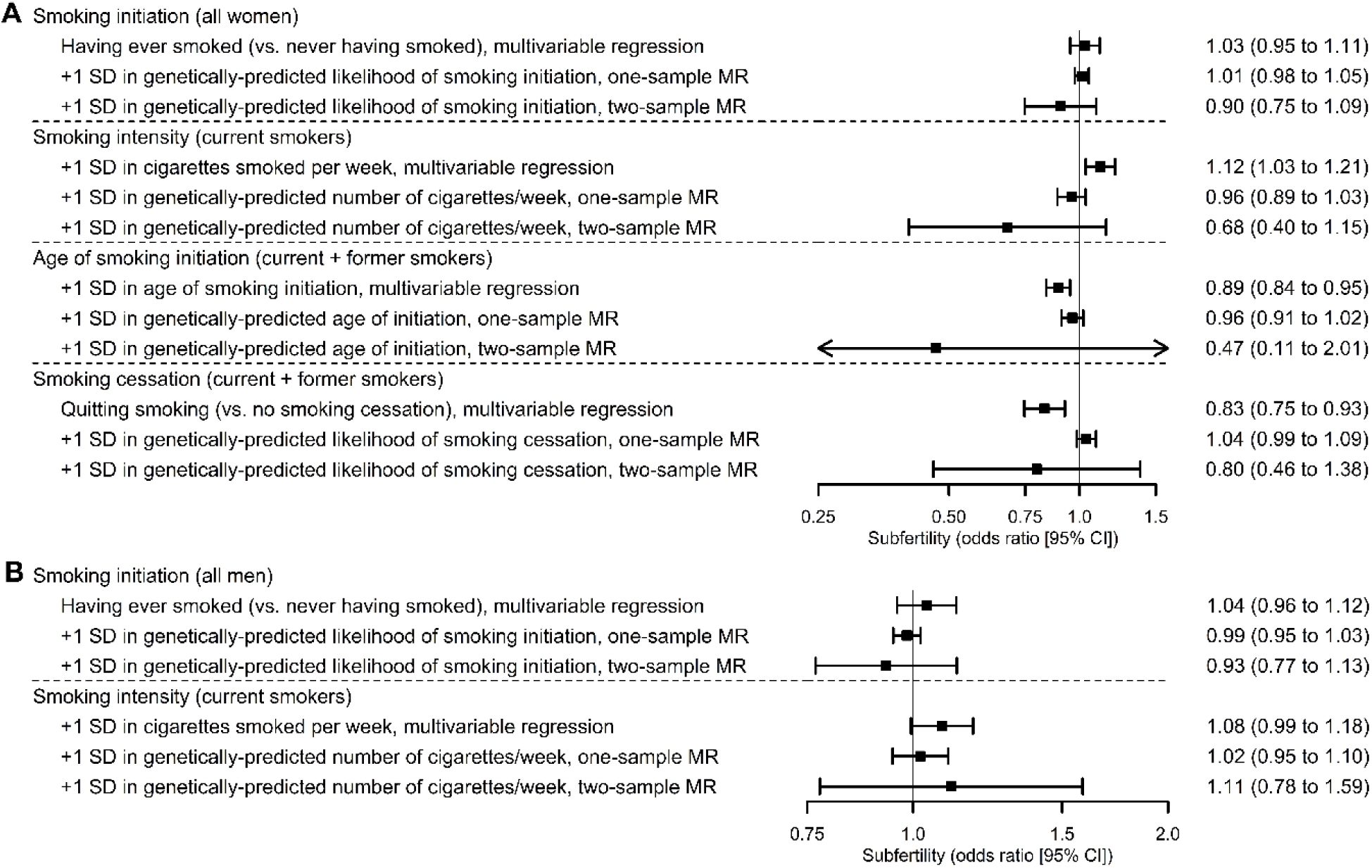
Association between smoking-related traits and subfertility in women (A) and men (B).

Results from the three methods for smoking initiation in men showed broadly similar patterns to those seen for women (**Figure 2B**). However, a linear association between higher smoking intensity and greater odds of subfertility was suggested in multivariable analyses (1 SD increase in the number of cigarettes smoked per week [+54 cigarettes/week]: OR 1.08, 95% CI 0.99 to 1.18; *p*-value_non-linearity_ = 0.123), attenuated when adjusting for the partner’s smoking intensity (OR 1.05, 95% CI 0.96 to 1.15; **Supplemental Table 2**). A similar but imprecise association was observed in two-sample MR (OR, 1.11, 95% CI 0.78 to 1.59), and close to the null results in one-sample MR (OR 1.02, 95% CI 0.95 to 1.10). It was not possible to complete multivariable regression and one-sample MR analyses in men, because detailed information on smoking was not obtained from fathers in MoBa. Nonetheless, in two-sample MR, close to the null, highly imprecise associations were found for age of smoking initiation (OR 0.95, 95% CI 0.19 to 4.68) and smoking cessation (OR 0.94, 95% CI 0.57 to 1.54).

### Sensitivity analyses

We observed associations between some smoking related GRSs and age (in women), education years (in both sexes) and body mass index (in both sexes) (**Supplemental Table 3**). Results of multivariable one-sample MR accounting for education and BMI and age-stratified analyses were consistent with the main analyses (**Supplemental Table 4**). No associations between the GRSs for smoking traits and subfertility were found in non-smokers (**Supplemental Table 5**).

Regarding two-sample MR additional methods, between SNP heterogeneity, potentially linked to horizontal pleiotropy, was observed for the genetic instrument for smoking intensity in women, as well as highly imprecise MR estimates (**Supplemental Table 6, Supplemental Figure 1**).

## DISCUSSION

We found close to the null associations between smoking initiation and subfertility, using confounder adjusted multivariable regression and MR in women and men. Regarding smoking heaviness, it was associated with greater subfertility odds in both sexes in multivariable regression. However, we found close to the null relationships in one-sample MR, a suggested clinically important inverse association in two-sample MR in women, and a substantial attenuation of the association in multivariable regression in men when adjusted for the partner’s smoking status. A similar pattern of association across the three methods was observed for age at initiation in women, whereas there were consistent inverse associations with smoking cessation between multivariable regression and two sample MR in women. Taken together, our findings do not provide strong evidence for effects of smoking on subfertility. However, results for two-sample MR were very imprecise, highlighting the need for large collaborative GWAS of subfertility, one-sample MR results may have been influenced by weak instrument and, despite numerous sensitivity analyses, we cannot rule out completely bias due to horizontal pleiotropy in MR analyses or residual confounding in the multivariable regression analyses.

A link between smoking and subfertility has been reported in human studies since the 1980s, particularly in women (5, 6, 44). Nevertheless, the American Society of Reproductive Medicine warned in 2018 of some methodological flaws in the available body of evidence including the risk of residual confounding, the small magnitude of the association, and the retrospective nature of most studies (14). We address these limitations here by assessing the association between several smoking-related traits and subfertility in a large prospective study comparing multivariable regression results to those form one-sample and two-sample MR, undertaking several sensitivity analyses to explore possible sources of bias. Our multivariable regression results were similar to previous studies, including point estimates that were close to the null for several smoking traits (6, 44). The magnitude of these relationships was modest, and they were generally consistent in direction to MR results, though with greater power and hence narrower confidence intervals. In particular, confidence intervals for estimates for smoking intensity and age at initiation (which are directly comparable) overlapped, suggesting the three sets of results were consistent with each other. It is possible that residual confounding due to poorer health status, worse diet quality, increased alcohol consumption, lower levels of physical activity, etc. (45-47) could explain the decreased fertility among smokers in multivariable analyses, with possible weak instruments in some of our one-sample MR analyses being biased towards these confounded results. Moreover, we could not rule out bias due to assortative mating completely (individuals who smoke are more likely to select a partner who also smokes (48), which may distort any potential association in couple-based outcomes such as fertility). Last, the genetic instruments for age of smoking initiation and smoking cessation were also weak. Although we used several approaches to minimize the likelihood of weak instrument bias, this could still have influenced findings, and in the two-sample MR in particular results were imprecise, suggesting low power to reject the null hypothesis (33, 38, 39, 49). Regarding men, we did not find any robust relationship between tobacco-related habits and subfertility across methods. These results are in line with previous prospective studies (10, 11).

Our work has some limitations. First, subfertility is a couple-dependent parameter that was reported by mothers in the cohort, and we could not determine the cause (female causes, male, or both). Second, MoBa is a pregnancy cohort, only including couples who were able to conceive. Further studies considering couples who never conceived (infertility) are needed. Subfertility is a less severe manifestation of infertility and, therefore, an association between smoking-related traits and an absolute incapacity to conceive may be possible. Third, our results may have been affected by selection bias as there was some differences between participants with and without genetic data. In addition, smokers are under-represented in the MoBa cohort relative to the whole Norwegian population (50). Nevertheless, associations between smoking and pregnancy outcomes (low birthweight, placental abruption, and stillbirth) proved unaffected by selection bias in prior studies when the MoBa cohort and the general Norwegian population were compared (50). Finally, our study population (adult women and men of a northern European ancestry who were capable of conceiving) restricts the generalizability. However, our work also presents several strengths. To our knowledge, this is the first prospective study assessing the relationship between smoking and subfertility using three complementary approaches affected by different sources of bias that have been thoroughly investigated and acknowledged. In addition, our study includes a relatively homogeneous population. This aspect minimizes the risk of confounding due to population stratification in MR, together with the adjustment for the first 10 principal components (41).

## CONCLUSIONS

We did not find robust evidence of an effect of smoking on subfertility. This may be due to a true lack of effect, weak genetic instruments, or other kinds of confounding. However, the comparison of different analytical approaches with complementary sources of bias has highlighted relevant limitations across all methods, and in particular highlights the needs for larger studies with information on subfertility.

## Data Availability

Consent given by the participants does not open for storage of data on an individual level in repositories or journals. Researchers who want access to datasets for replication should submit an application to. Access to datasets requires approval from the Regional Committee for Medical and Health Research Ethics in Norway and an agreement with MoBa.
Source data of the GWAS on smoking initiation, age of smoking initiation, smoking cessation, and smoking intensity) are available in the Supplemental Tables of the article by Liu M et al. (published in Nat Genet in 2019: https://www.nature.com/articles/s41588-018-0307-5#Sec14). Source data of the GWAS on education years are available in the Supplemental Tables of the article by Lee JJ et al. (published in Nat Genet in 2018: https://www.nature.com/articles/s41588-018-0147-3#Sec34). Finally, source data of the GWAS on BMI (Yengo L et al., Hum Mol Genet, 2018) are available in the GIANT Consortium website (https://portals.broadinstitute.org/collaboration/giant/index.php/GIANT_consortium_data_files#GWAS_Anthropometric_2015_BMI_Summary_Statistics).

## DATA AVAILABILITY STATEMENT

Consent given by the participants does not open for storage of data on an individual level in repositories or journals. Researchers who want access to datasets for replication should submit an application to. Access to datasets requires approval from the Regional Committee for Medical and Health Research Ethics in Norway and an agreement with MoBa.

Source data of the GWAS on smoking initiation, age of smoking initiation, smoking cessation, and smoking intensity) are available in the Supplemental Tables of the article by Liu M et al. (published in Nat Genet in 2019: https://www.nature.com/articles/s41588-018-0307-5#Sec14). Source data of the GWAS on education years are available in the Supplemental Tables of the article by Lee JJ et al. (published in Nat Genet in 2018: https://www.nature.com/articles/s41588-018-0147-3#Sec34). Finally, source data of the GWAS on BMI (Yengo L et al., Hum Mol Genet, 2018) are available in the GIANT Consortium website (https://portals.broadinstitute.org/collaboration/giant/index.php/GIANT_consortium_data_files#GWAS_Anthropometric_2015_BMI_Summary_Statistics).

## ACKNOWLEDGEMENTS

The MoBa Cohort Study is supported by the Norwegian Ministry of Health and Care Services and the Ministry of Education and Research. We thank all the participating families in Norway who take part in this ongoing cohort study, and those who contributed to the recruitment and the infrastructure of the cohort.

We thank the Norwegian Institute of Public Health for generating high-quality genomic data. This research is part of the HARVEST collaboration, supported by the Research Council of Norway (project reference: #229624). We also thank the NORMENT Centre for providing genotype data, funded by the Research Council of Norway (project reference: #223273), South East Norway Health Authority, and Stiftelsen Kristian Gerhard Jebsen. We further thank the Center for Diabetes Research (University of Bergen) for providing genotype information and performing quality control and imputation of the data in research projects funded by the European Research Council Advanced Grant SELECTionPREDISPOSED, Stiftelsen Kristian Gerhard Jebsen, the Trond Mohn Foundation, the Research Council of Norway, the Novo Nordisk Foundation, the University of Bergen, and the Western Norway Health Authority.

This work was performed on the TSD (Tjeneste for Sensitive Data) facilities, owned by the University of Oslo, operated and developed by the TSD service group at the University of Oslo, IT-Department.

This paper does not necessarily reflect the position or policy of the Norwegian Research Council.

