## Supplemental Materials for "Smoking and subfertility: multivariable regression and Mendelian randomization analyses in the Norwegian Mother, Father and Child Cohort Study"

**SUPPLEMENTAL METHODS**

*GWAS on subfertility in the MoBa cohort*

We assessed the relationship between the genotyped SNPs in the MoBa cohort after the quality control process (1) and subfertility by logistic regressions, in women and men separately. Any participant reporting a time-to-pregnancy ≥12 months or having undergone assisted reproductive technologies in any of their pregnancies was considered subfertile. The MoBa Genetics infrastructure is a collaborative research initiative arranged in five genotype batches of parent-offspring trios (1, 2). We evaluated the association between the genotyped SNPs and subfertility in each batch using Plink v1.9 (3) and lastly performed a sample size-weighted meta-analysis (random effects model) using the GWAMA software (4).

**SUPPLEMENTAL RESULTS**

*Robustness of genetic instruments for education years and body mass index*

The genetic instrument of education years was robust. Each one-unit increase in the GRS was associated with an increase of 0.028 education years in women (95% CI 0.026 to 0.030, *p* < 0.001, 4.11% of variance explained, *F*-statistic = 956) and 0.031 education years in men (95% CI 0.028 to 0.033, *p* < 0.001, 3.92% of variance explained, *F*-statistic = 837).

The genetic instrument of body mass index was also robust. In this case, each one-unit increase in the GRS was linked to an increase in body mass index of 0.044 kg/m^2^ in women (95% CI 0.041 to 0.046, *p* < 0.001, 5.65% of variance explained, *F*-statistic = 1,196) and 0.033 kg/m^2^ in men (95% CI 0.031 to 0.035, *p* < 0.001, 5.14% of variance, *F*-statistic = 1,036).

**SUPPLEMENTAL TABLES**

**Supplemental Table 1.** Comparison between participants with and without genotype information

|  | **Women** | | | **Men** | | |
| --- | --- | --- | --- | --- | --- | --- |
|  | Included | Non-included | *p*-value | Included | Non-included | *p*-value |
| Age (years), mean ± standard deviation | 30.3 ± 4.16 | 30.1 ± 4.76 | <0.001 | 32.7 ± 4.91 | 32.7 ± 5.61 | 0.690 |
| Education years,  mean ± standard deviation | 17.5 ± 3.12 | 16.9 ± 3.44 | <0.001 | 16.6 ± 3.50 | 15.9 ± 3.65 | <0.001 |
| Body mass index (kg/m^2^),  median (25^th^-75^th^ percentile) | 23.1  (21.2-25.9) | 23.1  (21.1-26.0) | 0.125 | 25.5  (23.7-27.7) | 25.4  (23.6-27.7) | <0.001 |
| Previous pregnancies,  *n* (%): |  |  | 0.379 |  |  | 0.045 |
| 0 | 13,016 (45.0%) | 31,718 (44.7%) |  | 12,408 (45.3%) | 32,326 (44.6%) |  |
| 1 or more | 15,902 (55.0%) | 39,236 (55.3%) |  | 14,979 (54.7%) | 40,159 (55.4%) |  |
| Subfertility, *n* (%) | 3,455 (11.9%) | 7,116 (9.97%) | <0.001 | 3,290 (12.0%) | 7,281 (9.99%) | <0.001 |
| Ever smokers (all participants),  *n* (%): | 13,575 (46.9%) | 36,642 (52.5%) | <0.001 | 13,353 (48.7%) | 34,013 (46.9%) | <0.001 |
| Cigarettes/week (current smokers), median (25^th^-75^th^ percentile) | 35.0  (7.00-70.0) | 49.0  (10.0-91.0) | <0.001 | 56.0  (7.00-105) | 70.0  (10.0-105) | <0.001 |

**Supplemental Table 2**. Association between smoking-related traits and subfertility in multivariable regressions

|  | Main analyses | | | Analyses adjusted for the corresponding trait of the partner | | Non-planned pregnancies in the reference group | |
| --- | --- | --- | --- | --- | --- | --- | --- |
|  | OR (95% CI) | *p*-value | Likelihood ratio test  (*p*-value) | OR (95% CI) | *p*-value | OR (95% CI) | *p*-value |
| **Women** | | | | | | | |
| Having ever smoked (vs. never having smoked) (all participants) | 1.03  (0.95 to 1.11) | 0.454 | - | 1.00  (0.92 to 1.09) | 0.948 | 0.99  (0.92 to 1.07) | 0.797 |
| +1 SD increment in the number of cigarettes smoked per week (current smokers) | 1.12  (1.03 to 1.21) | 0.005 | 0.970 | 1.10  (1.01 to 1.19) | 0.033 | 1.07  (0.99 to 1.15) | 0.088 |
| +1 SD increment in the age of smoking initiation (current + former smokers) | 0.89  (0.84 to 0.95) | <0.001 | 0.933 | - | - | 0.90  (0.85 to 0.95) | <0.001 |
| Quitting smoking (vs. no smoking cessation) (current + former smokers) | 0.83  (0.75 to 0.93) | <0.001 | - | - | - | 0.90  (0.81 to 1.00) | 0.049 |
| **Men** | | | | | | | |
| Having ever smoked (vs. never having smoked) (all participants) | 1.04  (0.96 to 1.12) | 0.366 | - | 1.02  (0.94 to 1.11) | 0.638 | 1.00  (0.93 to 1.09) | 0.940 |
| +1 SD increment in the number of cigarettes smoked per week (current smokers) | 1.08  (0.99 to 1.18) | 0.066 | 0.123 | 1.05  (0.96 to 1.15) | 0.274 | 1.04  (0.96 to 1.13) | 0.329 |

**Supplemental Table 3.** Linear associations between genetic risk scores and subfertility risk factors.

|  | Age (years) | Education (years) | Body mass index (kg/m^2^) | Previous pregnancies (n) |
| --- | --- | --- | --- | --- |
| **Women** | | | | |
| Smoking initiation GRS  (Δ1 SD) | -0.13 (-0.18 to -0.072)  (*p* < 0.001) | -0.22 (-0.26 to -0.17)  (*p* < 0.001) | 0.16 (0.10 to 0.22)  (*p* < 0.001) | 0.008 (-0.001 to 0.017)  (*p* = 0.087) |
| Smoking intensity GRS  (Δ1 SD) | -0.067 (-0.18 to 0.048)  (*p* = 0.253) | -0.026 (-0.12 to 0.069)  (*p* = 0.589) | -0.056 (-0.17 to 0.059)  (*p* = 0.338) | 0.003 (-0.017 to 0.023)  (*p* = 0.769) |
| Age of smoking initiation GRS  (Δ1 SD) | 0.082 (-0.008 to 0.17)  (*p* = 0.074) | 0.082 (0.009 to 0.15)  (*p* = 0.028) | -0.068 (-0.16 to 0.027)  (*p* = 0.162) | 0.003 (-0.012 to 0.017)  (*p* = 0.726) |
| Smoking cessation GRS  (Δ1 SD) | 0.029 (-0.052 to 0.11)  (*p* = 0.485) | 0.078 (0.014 to 0.14)  (*p* = 0.017) | 0.024 (-0.057 to 0.10)  (*p* = 0.565) | 0.010 (-0.012 to 0.014)  (*p* = 0.875) |
| **Men** | | | | |
| Smoking initiation GRS  (Δ1 SD) | -0.006 (-0.072 to 0.060)  (*p* = 0.866) | -0.25 (-0.29 to -0.20)  (*p* < 0.001) | 0.20 (0.16 to 0.25)  (*p* < 0.001) | -0.007 (-0.016 to 0.003)  (*p* = 0.152) |
| Smoking intensity GRS  (Δ1 SD) | 0.13 (-0.026 to 0.28)  (*p* = 0.103) | -0.091 (-0.20 to 0.014)  (*p* = 0.090) | -0.073 (-0.18 to 0.030)  (*p* = 0.163) | -0.004 (-0.024 to 0.017)  (*p* = 0.712) |

**Supplemental Table 4.** Multivariable and age-stratified one-sample MR analyses

|  | One-sample MR  (main analyses) | Multivariable MR (accounting for education years and body mass index) | MR: age of delivery  < median | MR: age of delivery  > median |
| --- | --- | --- | --- | --- |
| **Women** | | | | |
| Δ1 SD in the genetically predicted likelihood of smoking initiation  (all participants) | 1.01  (0.98 to 1.05) | 1.00  (0.94 to 1.05) | 1.06  (1.00 to 1.12) | 0.99  (0.95 to 1.04) |
| Δ1 SD in the genetically predicted number of cigarettes smoked/week (current smokers) | 0.96  (0.89 to 1.03) | 0.97  (0.90 to 1.05) | 0.97  (0.87 to 1.07) | 0.96  (0.86 to 1.06) |
| Δ1 SD in the genetically predicted age of smoking initiation  (current + former smokers) | 0.96  (0.91 to 1.02) | 0.99  (0.93 to 1.04) | 0.96  (0.89 to 1.05) | 0.96  (0.89 to 1.04) |
| Δ1 SD in the genetically predicted likelihood of smoking cessation  (current + former smokers) | 1.04  (0.99 to 1.09) | 1.11  (0.96 to 1.27) | 1.03  (0.96 to 1.11) | 1.04  (0.97 to 1.12) |
| **Men** | | | | |
| Δ1 SD in the genetically predicted likelihood of smoking initiation  (all participants) | 0.99  (0.95 to 1.03) | 0.95  (0.90 to 1.01) | 1.00  (0.94 to 1.06) | 0.98  (0.94 to 1.03) |
| Δ1 SD in the genetically predicted number of cigarettes smoked/week (current smokers) | 1.02  (0.95 to 1.10) | 0.94  (0.87 to 1.02) | 1.02  (0.91 to 1.14) | 1.01  (0.91 to 1.12) |

**Supplemental Table 5.** Associations between 1 SD increases in GRSs for smoking traits and subfertility in non-exposed participants (no-relevance sensitivity analyses)

| **Women** | |
| --- | --- |
| Δ1 SD in the genetically predicted  number of cigarettes smoked/week  (never + former smokers) | 0.96  (0.92 to 1.01) |
| Δ1 SD in the genetically predicted  age of smoking initiation  (never smokers) | 1.01  (0.96 to 1.06) |
| Δ1 SD in the genetically predicted  likelihood of smoking cessation  (never smokers) | 0.98  (0.93 to 1.04) |
| **Men** | |
| Δ1 SD in the genetically predicted  number of cigarettes smoked/week  (never + former smokers) | 0.99  (0.95 to 1.04) |

**Supplemental Table 6.** Estimates (odds ratios) of all two-sample Mendelian randomization methods plus indicators of horizontal pleiotropy and SNP heterogeneity.

|  | Inverse  variance  weighted | MR Egger | Weighted  median | Weighted  mode | MR-Robust Adjusted  Profile Score | Horizontal  pleiotropy  (MR Egger) | Cochran’s  Q | Rücker’s  Q’ |
| --- | --- | --- | --- | --- | --- | --- | --- | --- |
| **Women** | | | | | | | | |
| Δ1 SD in the genetically predicted likelihood of smoking initiation | 0.90  (0.75 to 1.09) | 1.12  (0.50 to 2.51) | 0.98  (0.74 to 1.29) | 1.12  (0.50 to 2.51) | 1.00  (0.83 to 1.19) | *p* = 0.595 | 239.91  (*p* = 0.995) | 239.62  (*p* = 0.995) |
| Δ1 SD in the genetically predicted number of cigarettes smoked/week | 0.68  (0.40 to 1.15) | 0.40  (0.11 to 1.46) | 0.60  (0.28 to 1.25) | 0.79  (0.31 to 2.06) | 0.67  (0.44 to 1.02) | *p* = 0.377 | 60.09  (*p* = 0.035) | 58.95  (*p* = 0.034) |
| Δ1 SD in the genetically predicted age of smoking initiation | 0.47  (0.11 to 2.01) | 1.91  (7×10^-4^ to 4950) | 0.28  (0.045 to 1.73) | 0.23  (0.021 to 2.67) | 0.68  (0.20 to 2.27) | *p* = 0.734 | 6.77  (*p* = 0.342) | 6.60  (*p* = 0.252) |
| Δ1 SD in the genetically predicted likelihood of smoking cessation | 0.80  (0.46 to 1.38) | 0.80  (0.16 to 3.97) | 1.40  (0.66 to 2.97) | 1.48  (0.54 to 4.02) | 0.85  (0.55 to 1.30) | *p* = 0.990 | 17.07  (*p* = 0.315) | 17.07  (*p* = 0.252) |
| **Men** | | | | | | | | |
| Δ1 SD in the genetically predicted likelihood of smoking initiation | 0.93  (0.77 to 1.13) | 0.91  (0.39 to 2.11) | 0.97  (0.74 to 1.29) | 1.82  (0.72 to 4.61) | 0.92  (0.76 to 1.10) | *p* = 0.956 | 225.47  (*p* = 1.000) | 225.47  (*p* = 0.999) |
| Δ1 SD in the genetically predicted number of cigarettes smoked/week | 1.11  (0.78 to 1.59) | 0.78  (0.41 to 1.47) | 0.82  (0.49 to 1.37) | 0.97  (0.59 to 1.60) | 1.04  (0.74 to 1.48) | *p* = 0.197 | 33.22  (*p* = 0.859) | 31.50  (*p* = 0.882) |
| Δ1 SD in the genetically predicted age of smoking initiation | 0.95  (0.19 to 4.68) | 4×10^-4^  (1×10^-7^ to 1.54) | 1.14  (0.16 to 8.13) | 5.71  (0.11 to 289) | 1.39  (0.39 to 4.88) | 0.120 | 7.34  (*p* = 0.29) | 3.84  (*p* = 0.573) |
| Δ1 SD in the genetically predicted likelihood of smoking cessation | 0.94  (0.57 to 1.54) | 1.72  (0.45 to 6.55) | 1.02  (0.52 to 2.01) | 1.09  (0.50 to 2.39) | 0.89  (0.58 to 1.37) | 0.353 | 5.47  (*p* = 0.987) | 4.55  (*p* = 0.991) |

**SUPPLEMENTAL FIGURES**

**Supplemental Figure 1.** Two-sample Mendelian randomization scatterplots for the associations of smoking initiation (A), age of smoking initiation (B), smoking cessation (C), and smoking intensity in women (D), and smoking initiation (E) and smoking intensity in men (F) with subfertility.

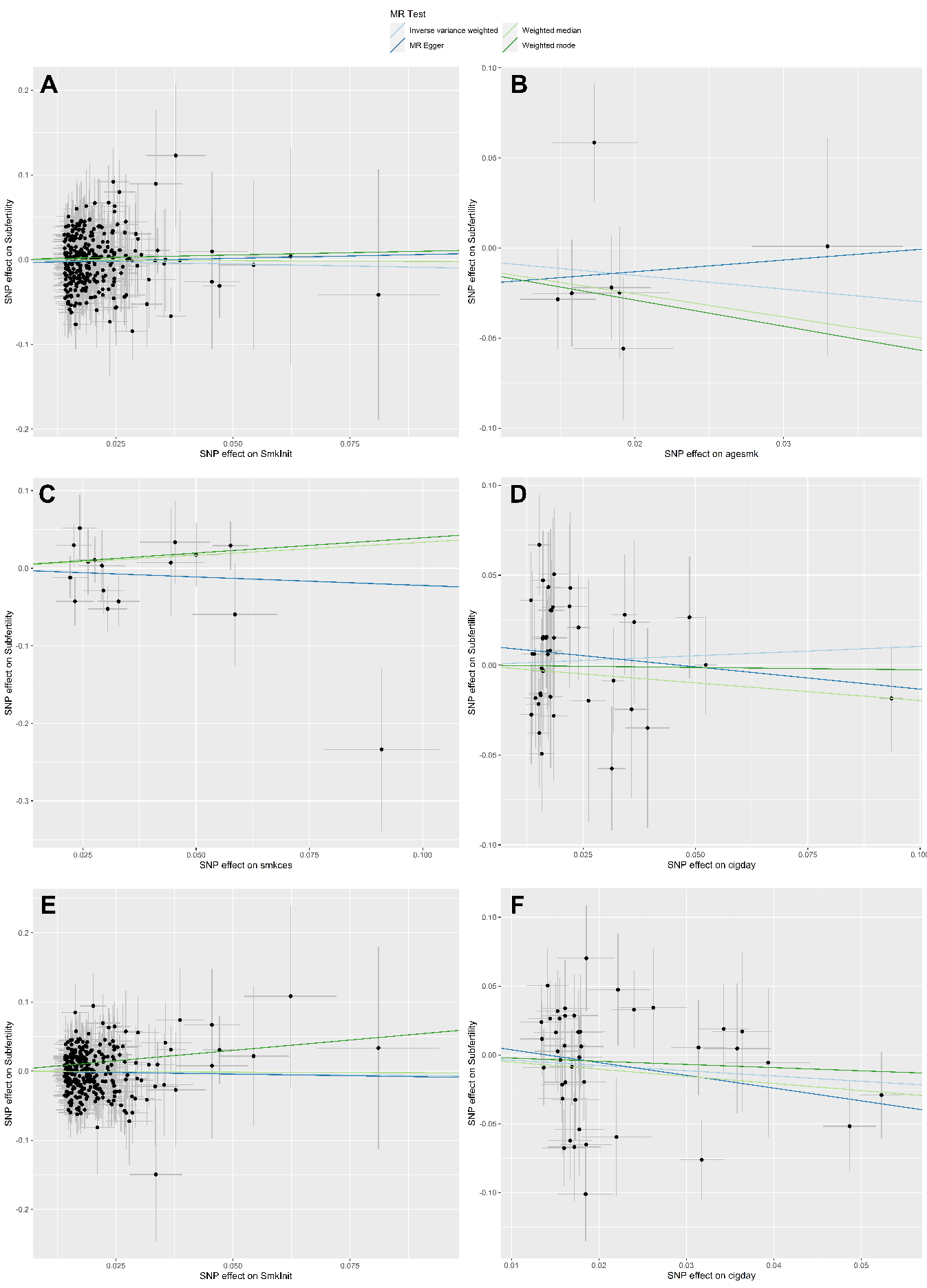
